# ReachUHC - A randomized controlled trial study protocol of mobile phone-based reminder and automatic renewal interventions to increase health insurance renewal rates in Kumasi, Ghana

**DOI:** 10.1101/2024.11.21.24317685

**Authors:** Laura Nübler, Mawumenyo Aku Kwawukume, Fati Ibrahim, Anne Neumann, Bernard Okoe-Boye, Vivian Addo-Cobbiah, Ellis Owusu-Dabo, Kofi Akohene Mensah, Verena Struckmann, Samuel Knauss, Julius Valentin Emmrich, Ruth Waitzberg, Enoch Affanyi, Orison Afflu, Joe Annor-Darkwah, Daniel Blankson, Francis Asenso-Boadi, Juliette Cazier, Jennifer Bencivenga, Carolina Pioch, Martin Siegel, Wilm Quentin, Daniel Opoku

**Author notes:** These three authors equally share last authorship (Martin Siegel, Wilm Quentin, Daniel Opoku).

## Abstract

**Background:** Since the Ghanaian National Health Insurance Scheme (NHIS) was introduced in 2004, coverage rates have remained low, despite affordable premiums and payment exemptions for minor, senior, poor and pregnant individuals. While 82% of the population have registered with the NHIS, many fail to complete the annual renewal and thus lose their coverage. A mobile renewal service introduced in 2018 simplified the previously cumbersome renewal procedure. Still, 40% of active member experience gaps in coverage in a given year, and 19% fail to renew at all. Baseline research suggests that forgetfulness is a major barrier to renewal.

**Methods:** 342,818 NHIS members from Kumasi will be randomized into the reminder, autorenewal or control groups. The reminder arm receives SMS prompts to complete the mobile renewal process and payment before expiration. The autorenewal arm is eligible to sign up for automatic renewal and give the NHIS permission to deduct the premium from their mobile money account, and will receive SMS prompt to do so. The intervention lasts 6 months. NHIS routine data will be used to evaluate the effect of the interventions on renewals. A follow-up survey household survey in Kumasi will evaluate additional aspects user experience.

**Discussion:** Improving insurance retention has the potential to substantially increase health insurance coverage rates, as 45% of the Ghanaians currently have expired insurance. Assessing these tools will identify enabling factors and barriers of the intervention and inform the transferability of the intervention to other health insurance systems in sub-Saharan African countries.

**Trial registration:** Pan African Clinical Trials Registry (PACTR)

## Introduction

### Background and rationale {6a}

Ghana implemented its National Health Insurance Scheme (NHIS) in 2004 in order to improve coverage and expand access to health care. While 82% of Ghanaians did register for NHIS membership at some point since the scheme’s inception, overall population coverage remains relatively low at about 52% of the population [1, 2]. Similar to health insurance schemes in other African countries, the NHIS relies on a system of annual re-enrolment of members [3]. This means that individuals have to actively renew their membership and, depending on their exemption status, may have to pay an annual premium and processing fee [2]. Members can renew their membership within 90 days before expiration. After the expiration date, there is a 90-day grace period in which the member can renew their membership without penalty and be able to instantly access care. After more than 90 days, the member can still renew but has a 30-day waiting period before insurance becomes active. This is done to discourage the likelihood of adverse selection of renewing health insurance only when needed. Children under 5, pregnant women and elderly over 70 are exempt from this waiting period.

Mobile technologies are increasingly used to support national health financing systems and contribute towards improved population coverage [4]. In Ghana too, the National Health Insurance Authority (NHIA) introduced a mobile renewal service in 2018. Until then, the insurance renewal process required personal attendance at an NHIS Office, and a number of studies investigating the reasons and determinants of non-registration and non-renewal found that long queues and waiting times required for in-person registration were significant deterrents to renewal, along with other factors, including dissatisfaction with the quality of healthcare and financial constraints [5, 6, 7, 8, 9]. Now, the mobile renewal service enables members to complete their renewal easily via mobile phones, using mobile money for payment of processing fees and premiums. The system was quickly accepted, with ∼70% of NHIS members renewing their insurance using the mobile renewal option [10], which has contributed to higher coverage rates among those who were previously less likely to be insured [2]. Despite this increase in convenience, around 50% of insurance periods are renewed either late or not at all each year. In a recent survey, members cited forgetfulness (19.8%), busy schedules (17.9%) and procrastination (15.6%) as reasons for non-renewal [11], which suggests that an intervention that reduces the logistical and organizational barriers may be effective in improving renewal rates.

Mobile phone-based reminders have been used in many countries and settings to improve adherence to medication or vaccination schedules, or to encourage timely preventative care or follow-up appointments [12, 13, 14]. Several studies have found text and voice message reminders to be effective in improving patient compliance and health outcomes [15, 16, 17, 18, 19, 14]. However, to the best of our knowledge, reminders have not been used to remind people to renew their health insurance in African countries, except as part of a broader intervention [20].

Additionally, automatic renewal of health insurance may potentially contribute to higher coverage rates [21]. Automatic renewal converts health insurance renewal into the default option upon a one-time opt-in. It can be considered a form of nudging that exploits a default-bias, whereby individuals are more likely to comply with a goal if it is made the default option and requires no additional action [22]. In the United States, automatic renewal has been found to lead to increasing health insurance coverage by reducing administrative barriers [21]. However, again, to the best of our knowledge, automatic renewal has not been used in an African context.

Therefore, the ReachUHC project has co-conceptualized an intervention consisting of regular renewal reminders for the NHIS, combined with an automatic renewal option to address the problem of coverage gaps. This intervention is to be introduced in Kumasi, Ghana. The main research questions are threefold: (1) What are the effects of renewal reminders and the automatic renewal service on coverage gaps and renewal rates?; (2) which factors are supporting intervention implementation and uptake, and what is the role of contextual factors?, and (3) is the intervention relevant and transferrable to other sub-Saharan African countries, specifically Kenya, Nigeria, Rwanda, Tanzania, and Uganda?

### Objectives {7}

The hypothesis of the study is that the ReachUHC intervention consisting of mobile phone-based reminders and an opt-in autorenewal option for NHIS renewal will increase renewal rates and reduce coverage gaps in Kumasi, Ghana. The overall aims of the project are to assess effectiveness of the intervention, analyze implementation processes, and study relevance and transferability of the intervention:

Objective 1: Evaluate the effectiveness of the ReachUHC intervention at increasing renewal rates and reducing coverage gaps.

Objective 2: Analyze user satisfaction, determinants of intervention uptake, and the need for intervention adjustment and improvement prior to national scale-up.

Objective 3: Explore relevance and transferability of the intervention to five African countries: Kenya, Nigeria, Rwanda, Tanzania, and Uganda

### Trial design {8}

This trial is a single-site RCT. NHIS members in Kumasi will be cluster-randomized by phone number into a parallel-group multi-arm superiority trial with a 1:1 allocation ratio to evaluate the effectiveness of renewal reminders and the autorenewal platform on active and recently expired members.

### Methods: Participants, interventions and outcomes Study setting {9}

Our study is conducted in the city of Kumasi, Ghana’s most populous city with 3.5 million inhabitants. We draw our study population from the 1.37 million NHIS members living in all of Kumasi’s Sub-Metropolitan Districts (Asawase, Asokwa, Bantama, Bosomtwe-Atwima-Kwanwoma, Manhyia and Subin), and in one of its major suburban districts (Ejisu-Juaben). We chose Kumasi as our study site as it provides a diverse population in a populous, geographically contained area.

### Eligibility criteria {10}

1. Participants must have active or recently expired NHIS membership at the starting date of the intervention. If expired, the expiration date of their insurance must be no older than 1 October 2022
2. Participants must be identified as residents of the Kumasi districts either through the location in their member file in the NHIS database or through the survey or phone number verification exercise conducted in 2023.
3. Participants must have a phone number which is registered as the primary phone number for no more than 6 NHIS members residing in Kumasi.

During a phone number verification exercise conducted by the NHIA in September 2023, a group of 64,646 NHIS members in Kumasi was surveyed in order to verify or update their contact details and mobile phone number. All individuals who were surveyed as part of this exercise, the “verified number sample”, are recruited as participants. The individuals surveyed during the preliminary implementation research survey in February and March 2023 were also included in this verified sample (if they were NHIS members and fit the eligibility criteria). Furthermore, any individual identified as a Kumasi resident in the database who was not included in the verification exercise and fulfills the inclusion criteria is eligible to be randomly selected into the “unverified number sample”, which forms the remainder of the participants. The verified sample consists of a group of 61,097 eligible individuals, the unverified sample consists of 281,281 eligible members which were randomly drawn from all eligible Kumasi residents in the NHIS database.

### Who will take informed consent? {26a}

A waiver for informed consent has been granted from the Kwame Nkrumah University of Science and Technology Committee on Human Research, Publication and Ethics. We argue that informed consent is not needed in this case according to the requirements set forth by the Council for International Organizations of Medical Sciences (CIOMS), World Health Organization (WHO), and the US Department of Health and Human Services (HHS) that

- risk to participants is minimal [23]
- the research has important social value [23]
- the research could not be practicably carried out without the waiver [23, 24]
- the study is conducted by public health authorities and is designed to study a public service program, its procedures for obtaining benefits, and possible changes in those procedures [24]

Members will not be prospectively informed of their participation in the trial and will not be asked to consent to receiving SMS messages. However, the sign-up to the autorenewal platform is voluntary, and NHIS members who do not wish to receive further SMS communications from the NHIA can opt out any time via USSD menu. We consider this approach appropriate, as consent to receive SMS communications from the NHIA was given implicitly by all individuals who registered for membership with the NHIS. Since residents of Ghana commonly receive text message reminders for different subscriptions and memberships, an SMS reminder for health insurance renewal will not be unusually intrusive or present a significant burden. The sign-up for the autorenewal service is voluntary, and users may unsubscribe from it through a simple process in the USSD menu at any time. Furthermore, users are prompted to enter a PIN to authorize payment before funds can be deducted from their mobile money account, so consent for the payment will also be obtained. To the best of our knowledge, there is no conceivable risk or adverse effect which can be expected to result from these services. As we evaluate the participants using only anonymized routine data in the primary study, no further consent provisions will be required.

### Additional consent provisions for collection and use of participant data and biological specimens {26b}

For both the quantitative follow-up survey and the qualitative scale-up interviews, the administrators explain a prepared statement on informed consent (translated into applicable local languages) to all selected participants before administering the questionnaire and obtained a signature (or fingerprint) from the participant on the informed consent form. The statement of informed consent 1) provides a description of study goals and survey procedure, 2) asserts voluntary nature of participation and the right to withdraw anytime, 3) provides information over confidentiality, privacy and intended use of survey data, and 4) assures that non-participation will not result in any disadvantages for the potential participant. After the statement has been explained, survey administrators offer to answer any questions that the potential participant may have. After all outstanding questions have been answered, the potential participant can choose to sign and date the form and enrol in the survey, or not sign the form and not enrol in the survey. The survey administrator also provides name, signature and date on the form as a witness to the consent given.

### Interventions

#### Explanation for the choice of comparators {6b}

We break down our samples into 3 groups based on their membership status and expiration dates. Group 1 contains all individuals with active insurance and an expiration date during the intervention period. Group 2 contains all individuals with a currently expired membership and expiration date between 1 October 2022 and the end of the pre-intervention period. Group 3 contains all members with active insurance and an expiration date after the planned end date of our intervention period.

We do this because expired members require different reminders and autorenewal messages than active members, and because those with a renewal date during the intervention phase receive messages around their expiration dates which individuals with renewal date outside of this time frame cannot receive. Furthermore, we expect that individuals with expired insurance may react differently than individuals with active insurance, and that those with more imminent renewal dates may respond differently than those whose renewal dates are still far away. We randomly allocate treatment within each group.

Due to the randomization, the treatments and control within each expiration date group (1, 2, 3) arms only differ by the treatment status. Individuals in the control arms will not receive SMS prompts for renewal or autorenewal, but will otherwise receive usual communications from the NHIA which are not related to this trial. Only participants assigned to be treated in the autorenewal arm will be able to sign up for the autorenewal platform as primary users, but will be able to add other individuals (typically dependents and other family members) to their renewal plan and pay for their renewals. All other individuals will be able to access the USSD Menu of the autorenewal platform through the short-code, but will not be able to sign up for the service. Instead, they will be shown an error message informing them that this platform is still undergoing testing and may be made available at a later date.

#### Intervention description {11a}

##### Treatment 1: Reminders

###### Arm 1R

Participants in the reminder arm with expiration dates during the intervention period will be targeted with the following SMS messages sent to their phone number at the specified intervals before their individual expiration dates.

| Message # | 1 | 2 | 3 | 4 | 5 | 6 |
| --- | --- | --- | --- | --- | --- | --- |
| $t_x$ | $t_{-28}$ | $t_{-7}$ | $t_0$ | $t_{+30}$ | $t_{+60}$ | $t_{+89}$ |
| Condition | Membership has not been renewed by $t_x$ | | | | | |
| Message | Dear<br>Firstname<br>Lastname,<br>Your<br>NHIS<br>card will<br>expire in<br>*4 weeks*.<br>You can<br>renew<br>using the | Dear<br>Firstname<br>Lastname,<br>Your NHIS<br>card will<br>expire in *1 week*. You<br>can renew<br>using the<br>MyNHIS<br>app, *929# | Dear<br>Firstname<br>Lastname,<br>Your NHIS<br>card expires<br>*TODAY!*<br>You can<br>renew using<br>the MyNHIS<br>app, *929#<br>or go to the | Dear<br>Firstname<br>Lastname,<br>Your NHIS<br>card is<br>expired. You<br>can renew<br>within 60<br>days to avoid<br>a 1-month<br>wait. Use | Dear<br>Firstname<br>Lastname,<br>Your NHIS<br>card is<br>expired. You<br>can renew<br>within 30<br>days to avoid<br>a 1-month<br>wait. Use | Dear<br>Firstname<br>Lastname,<br>Your NHIS<br>card is<br>expired. You<br>can still<br>renew today<br>to avoid a 1-month wait.<br>Use |
|  | MyNHIS app, *929# or go to the nearest NHIA district office. | or go to the nearest NHIA district office. | nearest NHIA district office. | MyNHIS app, *929# or go to the NHIA district office. | MyNHIS app, *929# or go to the NHIA district office. | MyNHIS app, *929# or go to the NHIA district office. |

##### Arm 2R

Members with already expired insurance only receive the following renewal reminder twice.

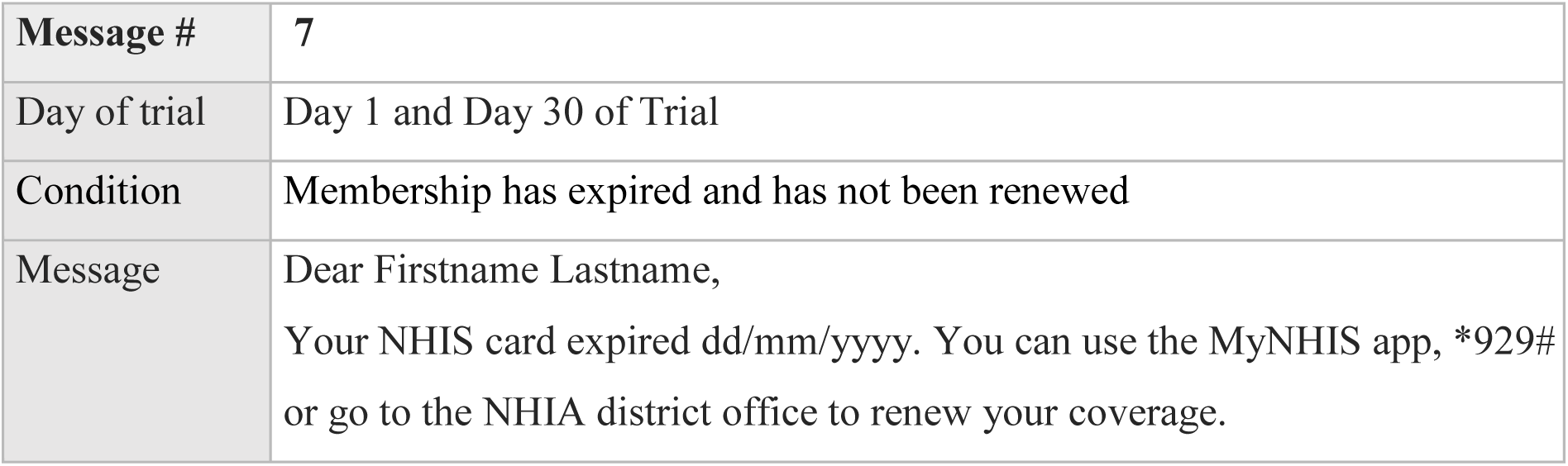

|  |  |
| --- | --- |
| Message # | 7 |
| Day of trial | Day 1 and Day 30 of Trial |
| Condition | Membership has expired and has not been renewed |
| Message | Dear Firstname Lastname,<br>Your NHIS card expired dd/mm/yyyy. You can use the MyNHIS app, *929# or go to the NHIA district office to renew your coverage. |

### Treatment 2: Autorenewal

All participants in these arms will receive Message 1 on Day 1 and Day 30 of trial (if they have not yet signed up). By dialing the short-code *929*20#, participants will be directed to a USSD-Menu where they can opt into the autorenewal service by providing their member number and then authorizing the automatic payment from their MoMo account to the NHIS when the renewal become due. A participant can also add the member number of any other individual (e.g. spouse or child) who they wish to add to the renewal plan.

### Arm 1A, 2A, 3A

Once signed up, individuals in arm 1A will receive messages 2 and 3 on the indicated days, provided that membership has not yet been renewed otherwise. The automatic renewal is triggered 30 days before the expiration date. If the expiration date is less than 30 days away or the membership is already expired, it will be triggered the day after sign-up. If no payment or further documentation is required, insurance is renewed immediately. If a payment is required, it will be debited to the MoMo account of the participant. If payment is successful, the insurance is renewed. If payment is unsuccessful, the member is informed and the renewal will be triggered in the 3 subsequent days again. After that, the member will receive a final message suggesting they attempt renewal via one of the other platforms.

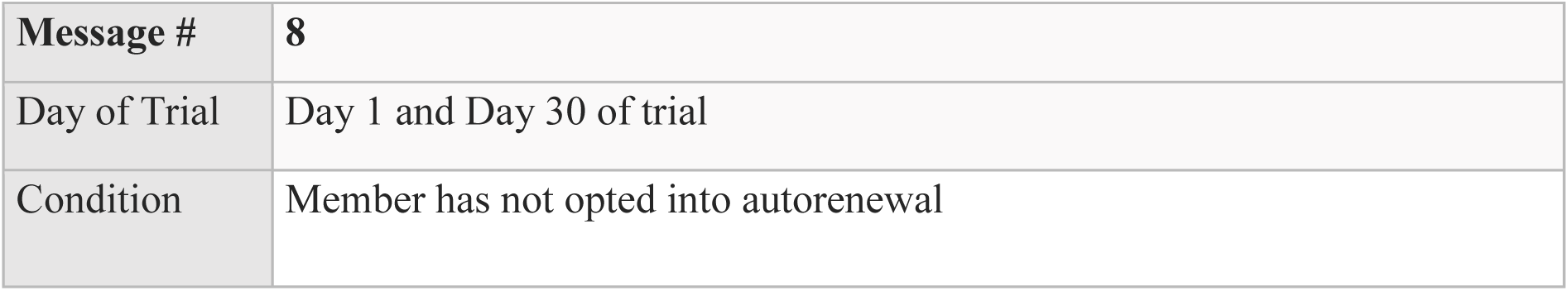
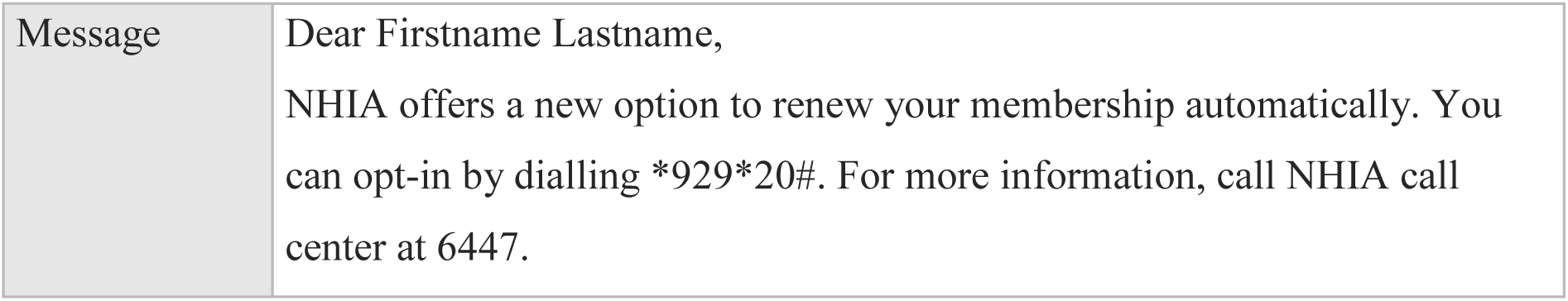

Arm 1A, 2A, 3A
|  |  |
| --- | --- |
| Message # | 8 |
| Day of Trial | Day 1 and Day 30 of trial |
| Condition | Member has not opted into autorenewal |
| Message | Dear Firstname Lastname,<br>NHIA offers a new option to renew your membership automatically. You can opt-in by dialling *929*20#. For more information, call NHIA call center at 6447. |

### Arm 1A

**Arm 1A, 2A, 3A -** If expiration is 30 days or less away on day of opt-in (or already expired)

Arm 1A
| Message # | 9 | 10 | 11 | 12 |
| --- | --- | --- | --- | --- |
| t<sub>x</sub> | t-90, t-60 | t-31 | t-30, t-29, t-28, | t-27 |
| Condition | Member has opted into autorenewal. Active membership has not been renewed. The member is required to pay premium or processing fee. |  | Autorenewal payment has failed | Autorenewal payment has failed 4 times |
| Message | Dear Firstname Lastname,<br>your NHIS insurance automatically renews on dd/mm/yyyy.<br>Please make sure you have xx cedis in your MoMo account. | Dear Firstname Lastname,<br>your NHIS insurance automatically renews in one day.<br>Please make sure you have xx cedis in your MoMo account. | Dear Firstname Lastname,<br>unfortunately, your membership could not be renewed. We will try again tomorrow. Please make sure you have xx cedis in your MoMo account. | Dear Firstname Lastname,<br>your membership could not be renewed automatically.<br>Please use MyNHIS app, *929# or go to the nearest NHIA district office. |
|  | (Dial *929*20# to change your settings) | (Dial *929*20# to change your settings) | (Dial *929*20# to change your settings) |  |

**Arm 1A, 2A, 3A** - If expiration is 30 days or less away on day of opt-in (or already expired)
| Message # | 11 | 12 |
| --- | --- | --- |
| Condition | Autorenewal payment has failed | Renewal has failed 4 times |
| Day of sign-up/opt-in (s<sub>0</sub>) | S+1, S+2, S+3 | S+4 |
| Message | Dear Firstname Lastname, unfortunately, your membership could not be renewed. We will try again tomorrow. Please make sure you have xx cedis in your MoMo account.<br>(Dial *929*20# to change your settings) | Dear Firstname Lastname, your membership could not be renewed automatically.<br>Please use MyNHIS app, *929# or go to the nearest NHIA district office. |

### Criteria for discontinuing or modifying allocated interventions {11b}

Participants can opt out from receiving SMS messages from the NHIA and not receive further communication at any time. Opt-out requests and sign-up rates to the autorenewal system will be monitored intermittently as indicators of protocol adherence and as warning signs of any unforeseen problems. Changes to the protocol may be made if adverse effects on participants or technological faults which threaten the validity of the trial are discovered.

### Strategies to improve adherence to interventions {11c}

In order to reduce technological and logistical barriers, the NHIS Helpline and local NHIA office staff will be trained to answer questions about the autorenewal system and assist with enrolment if needed.

Individuals with a phone number shared by more than 5 people are excluded from the trial in order to prevent excessive messaging to a single number, which would increase likelihood of opting out.

### Relevant concomitant care permitted or prohibited during the trial {11d}

There are no requirements or restrictions on care or other interventions for individuals during the duration of this trial.

### Provisions for post-trial care {30}

We do not expect any potential for harm from reminder messages or the automatic renewal option. However, should any problem occur, the NHIS members support services will be able to assist with any potential issues.

### Outcomes {12}

Primary outcomes

| **Outcomes** | **Analysis Metric** | **Method of aggregation** | **Time point** |
| --- | --- | --- | --- |
| Renewal status<br>(renewal by expiration date, renewal without coverage gap, any renewal) | Absolute difference and Odds Ratio between treatment and control, between expiration date groups | Proportion succeeding in renewing insurance coverage before expiration | End of intervention period<br>Day 180 |
| Coverage gap | Absolute and relative | Mean length in days/duration | End of intervention period |
|  | difference, as well as hazard ratios, between treatment and control, between expiration date groups | until renewal | Day 180 |

Secondary outcomes

| **Outcomes** | **Analysis Metric** | **Method of aggregation** | **Time point** |
| --- | --- | --- | --- |
| Acceptance of autorenewal option | Odds Ratio between treatment and control, | Proportion signing up for autorenewal | End of intervention period |
| Successful renewal via autorenewal | Odds Ratio between treatment and control | Proportion of autorenewal set-ups resulting in a successful renewal | End of intervention period |
| User experience and acceptability of intervention | Agreement with questions regarding usefulness and usability of intervention | Proportion of individuals agreeing with statements | Follow-Up Survey<br>Post-Intervention period |

### Participant timeline {13}

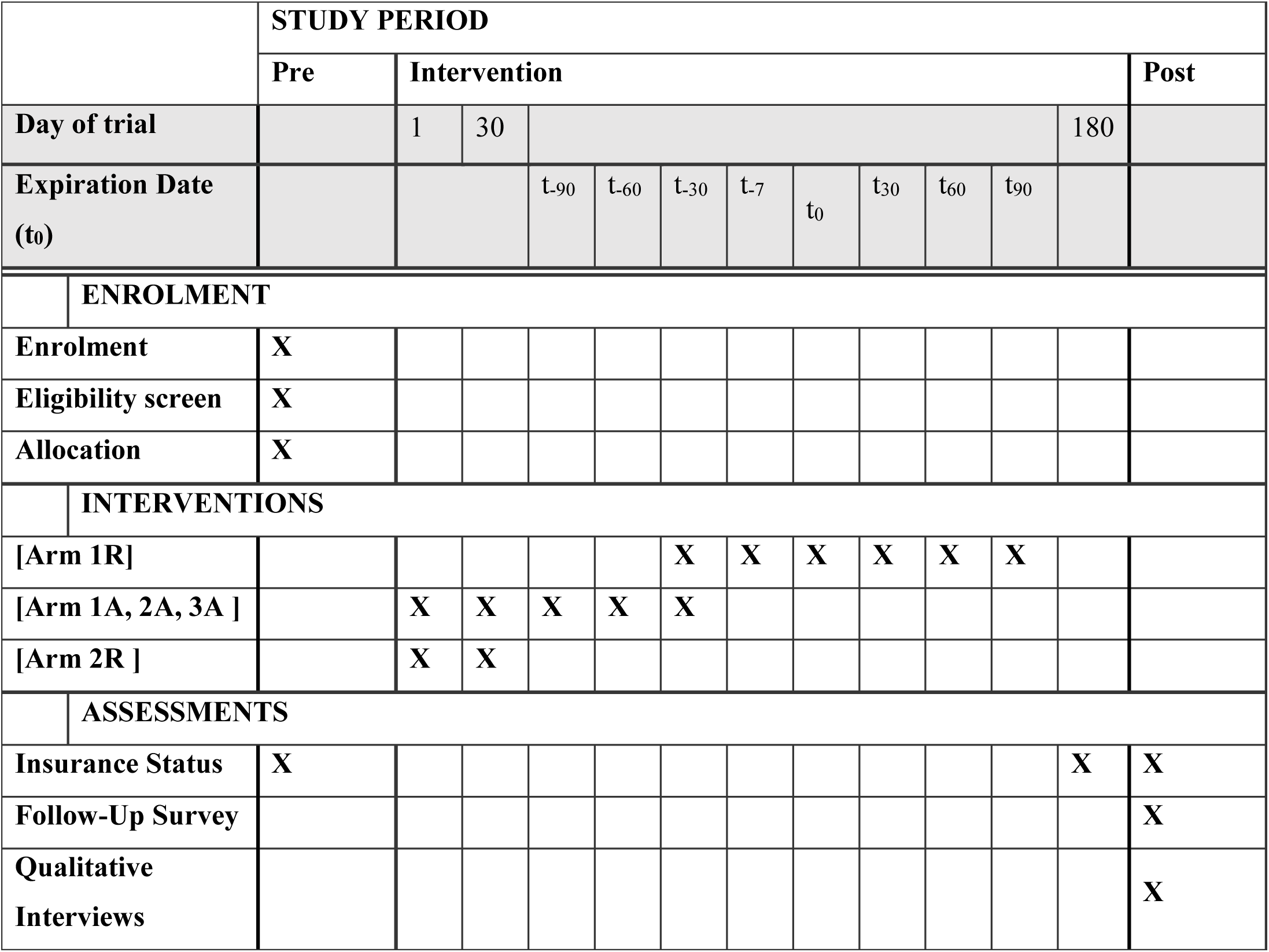

#### Sample size {14}

We compute sample size estimations for a binary-outcome, parallel group superiority trial assuming 95% significance (α=0.05), 80% power (β=.20) and a normal distribution [25, 26]. In order to account for the clustered randomization, we inflate the sample size estimate with the design effect coefficient [(1+ (n-1) ρ], where n is the cluster size and ρ is the intraclass correlation coefficient [27, 28]. We assume an intracluster correlation coefficient of ρ=0.2, an average cluster size of n=1,3 for the verified sample, and n=1,7 for the unverified number sample based on our estimates of sample characteristics. We conservatively assume an effect size of 10% based on systematic reviews of SMS-based reminder systems in health care settings [29, 15, 16, 17, 18, 19, 14, 30]. This gives us a requirement of 2,530 individuals per arm. Since the phone number validation exercise showed that 48% of NHIS members did not have their current, up-to-date phone number on file, we must assume that only a fraction of SMS-reminders in the unverified population will reach the intended target, so we can expect smaller effect sizes for the unverified number sample. If unintended spill-overs occur, this could further attenuate effect size. As a result, we chose to randomly select 30,000 phone number clusters into each arm for the unverified number sample. This is the maximum sample size feasible due to budget constraints. With this sample size, an effect size as small as 3.46% would be detectable in the unverified number sample.

#### Recruitment {15}

Recruitment for the intervention study consists of selecting members to target with SMS messages from the NHIS database and the verified number sample. The phone number verification exercise which was conducted in September 2023 will ensure that a high percentage of targeted individuals can be successfully contacted and thereby enrolled. The remaining participants are recruited and enrolled automatically from the NHIS database right before the start of the intervention, which ensures that a sufficient number of individuals will be successfully contacted, despite the limitations stemming from outdated or shared phone numbers.

For the quantitative follow-up survey, we recruit the same 280 participants which had already been surveyed as part of a preliminary implementation survey in February-March 2023 and which are registered NHIA members meeting the eligibility criteria of this study.For the transferability and international scale-up research, we recruit participants in the selected target countries Kenya, Tanzania, Uganda, and Nigeria. We adopt systematic reviews of both peer-reviewed and grey literature, as well as a qualitative approach to collect data using individual semi-structured interviews (with stakeholders) as well as focus groups (with informal sector workers). The stakeholders are sampled purposively [31] from institutions involved in the development and implementation of the existing digital technologies for health financing (DTHF). We chose purposive sampling as the method as it is appropriate for identifying and selecting individuals or groups of individuals who are especially knowledgeable about or experienced in the subject matter [32]. Stakeholders who respond to our request and are available at the time of our study will be interviewed. We will use community recruitment for the informal sector workers. We will purposefully select participants and (e.g., home visits, door-to-door, friends or family, markets) based on high percentages of informal workers in particular districts of the capital city in each of the countries. Enrolment is not limited by geographic location. The strata used for purposive sampling include the variables (stakeholders = age, gender, years working in the institution, role; informal sector workers = age, gender, insurance status, place of residence). Informal workers will be recruited by convenience sampling techniques based on their availability and willingness to take part in the study. Sampling will continue until saturation [33], meaning no new themes or codes emerge from additional interviews and focus group discussions.

#### Assignment of interventions: allocation

##### Sequence generation {16a}

To select the unverified sample, we randomly draw 240,000 phone number clusters from a total of 1,128,542 eligible individuals in the NHIS database who were not already identified during the verification exercise. As phone numbers are frequently re-assigned by the phone companies or passed on to relatives and friends, all unverified individuals with a phone number on file that is shared with someone in the verified sample are excluded to prevent spillovers. We then split our verified and unverified samples into the expiration date groups 1, 2 and 3 as described above. In order to prevent spill-overs between the groups, individuals in the auxiliary groups (2, 3) who share a phone number with an individual in the main group (1) are also removed from the sampling population before randomization. Group 1 is cluster-randomized by phone number into the arms 1R (reminders), 1A (autorenewal) and 1C (control) with 1:1:1 proportion. Group 2 are cluster-randomized by phone number into the arms 2R (reminders), 2A (autorenewal) and 2C (control) with 1:1:1 proportion. Group 3 is cluster-randomized by phone number into arms 3A (autorenewal) and 3C (control) with 1:1 proportion. The randomizr command executed in Stata (15.1) uses a computer-generated sequence to randomly assign treatment by assigning a number between 0 and 2 (for groups 1-2) and 0 and 1 (for group 3) to each individual phone number cluster. The researchers pass on the allocation list with the anonymous unique identifiers to the technical implementation team, which executes the sending of the messages according to this allocation. Researchers do not remain blinded after the allocation has been completed in order to begin monitoring technical functioning and outcomes.

**Figure 1.**
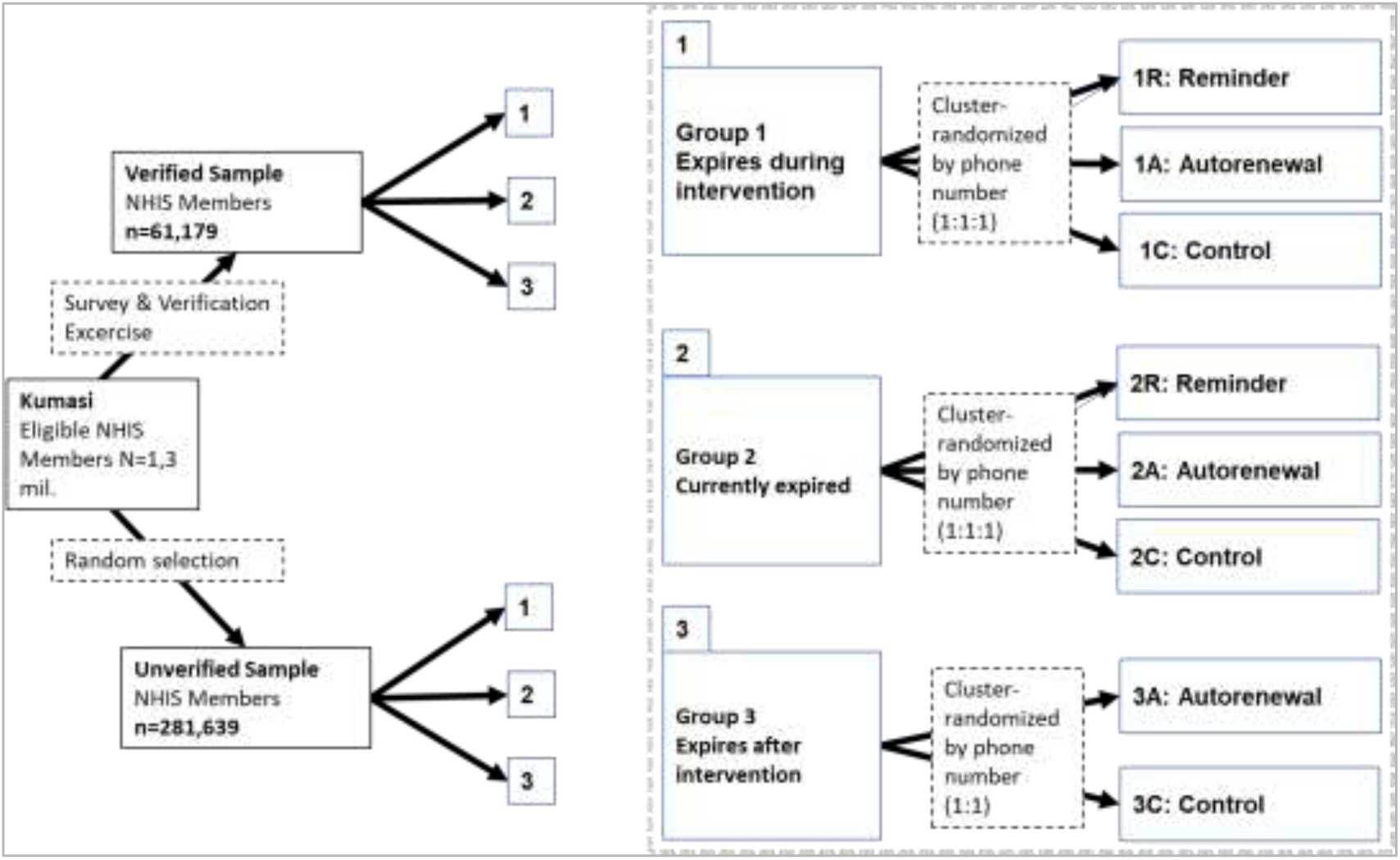
Sampling and Allocation Strategy

#### Concealment mechanism {16b}

The randomizr command conceals the sequence.

#### Implementation {16c}

The command randomizr is used by one of the researchers to employ the allocation sequence in each group of participants. Once selected, a participant is automatically enrolled.

### Assignment of interventions: Blinding

#### Who will be blinded {17a}

No blinding is done after assignment.

#### Procedure for unblinding if needed {17b}

N/A.

### Data collection and management

#### Plans for assessment and collection of outcomes {18a}

Anonymized routine data of all trial participants is extracted from the NHIS database, and shared with the researchers. This contains the renewal transaction data of NHIS members (date of transaction, beginning and end of insurance period, fees paid, platform used for transaction), intervention status (trial arm, autorenewal registration status, opt-out status) and user information (unique identifying number, age, gender, marital status, payment category) for each participant. This data is used to assess rates of insurance renewal and of autorenewal enrolment across treatment arms and demographic subgroups.

In addition, a randomly selected sample of 280 households in Kumasi will be surveyed as part of the quantitative follow-up survey assessing user experience, acceptability and usability of the reminder and autorenewal service. These households were previously surveyed in 2023 as part of a preliminary implementation and feasibility study, and will be surveyed again after the intervention is completed. The data will be collected by trained research assistants through face-to-face interviews. The quantitative follow-up survey will collect sociodemographic characteristics of households and individuals, health insurance coverage status, satisfaction with the reminders and autorenewal option.

For the qualitative scale-up interviews, data will be collected using a semi-structured approach based on interview guides. The interview guide used for data collection will be prepared based on existing literature and the research objectives of the study.

#### Plans to promote participant retention and complete follow-up {18b}

Participants can only deviate from the treatment protocol by opting out of receiving SMS messages. However, they can still be observed in the routine data after opting out, so will automatically be followed up on.

### Data management {19}

Routine data is captured automatically in the NHIS data management system. All data is anonymized being sent to researchers via a secure, password-protected cloud server, and under a non-disclosure agreement.

Data for the quantitative follow-up survey data is captured using REDCap, a secure, web-based electronic data capture system. It will be managed at the KNUST and hosted by their server. Only staff listed in the delegation log is given the unique individual password to access the internet-based data management system. The data is exported from the REDCap software into excel to validate the data and check for missing data. Submitted surveys is validated daily to ensure mistakes are seen early enough for corrections to be made and to ensure the number of interviews done tally with the data submitted to prevent data loss and ensure data uniformity. Researchers keep anonymized routine and survey data only on password-protected computers which comply with the university’s security standards. Both routine and survey data undergo plausibility checks for range and distribution of values before analysis.

Data for the qualitative scale-up interviews is collected anonymously and treated confidentially. Audio-recorded interviews will be transcribed verbatim. The data is stored on secure server at the KNUST and transferred to consortium partners in accordance with established data transfer agreement, and is only published in aggregated form in publications by the study team. The data is used for research projects and possible scientific publications of the participating project team.

Participants contact details (name, e-mail address, address) are collected and stored in encrypted form, i.e. neither name, initials nor address etc. appear in the encryption code. The data is only used to contact them for project coordination purposes and will be deleted at the end of 2024. The audio recordings are converted into text (transcribed) and then deleted. The transcription is carried out by the KNUST partner university and the text is anonymized. This means that information that allows conclusions to be drawn about participants is blacked out or changed accordingly.

### Confidentiality {27}

Survey participants will be assured of the confidentiality of the survey process before they give consent. The survey team will ensure a private setting for interviews to be conducted. Any data passed on to researchers will only be identified by a unique personal number to ensure anonymity. Only data administrators will have access to identifying information.

### Plans for collection, laboratory evaluation and storage of biological specimens for genetic or molecular analysis in this trial/future use {33}

N/A

## Statistical methods

### Statistical methods for primary and secondary outcomes {20a}

Descriptive statistics for each treatment arm at baseline will summarize sociodemographic characteristics as means, percentages or counts with standard deviations. The primary outcomes will be assessed as the difference between each treatment and the control group, comparing the difference between those who received renewal reminders (1R and 2R), autorenewal prompts (1A and 3A), or nothing (1C and 2C). As we expect active and expired members to respond differently to the messages, each treatment arm is also compared to its corresponding control arm within each group (i.e. 1R, 1A and 1C will be compared to each other, then 2R, 2A and 2C will be compared to each other separately). The main outcome is the proportion of successful renewals in each group.

Effect sizes will be estimated with a logistic regression of treatment arm on insurance outcome, and reported as both absolute differences and ORs with significance levels. To assess whether reminders resulted in less or in shorter coverage gaps, we will conduct a duration analysis, comparing time-to-renewal between the treatment arms using Kaplan-Meier curves and Weibull regressions. All effect sizes will represent an intent-to-treat (ITT) effect, as we cannot be certain if a treatment reached the intended recipient. The secondary outcomes will be assessed using descriptive statistics and logistic regressions in order to compare the likelihood of signing up for autorenewal, and the likelihood of a successful renewal occurring, conditional on having signed up for autorenewal, across the different expiration date groups. The results will be reported as absolute differences and odds ratios.

The quantitative follow-up survey data will be analysed using descriptive statistics showing the distribution to responses to answers across the Likert scale, and logistic regressions of sociodemographic factors on reported outcomes.

The qualitative scale-up interview data will undergo a thematic analysis with ATLAS.ti 22. The interview transcripts will be coded independently to validate and confirm the coding strategy. The codes will be sorted into themes and subthemes and refined into a narrative. A combination of deductive and inductive methods will be applied for data analysis. Deductive reasoning will be guided specifically by using the Predisposing, Need, and Enabling Framework and other existing literature on the factors influencing the implementation and potential use of mobile technology [9, 34, 35].

### Interim analyses {21b}

Opt-out requests and autorenewal sign-ups will be monitored to assess error-free functioning of the reminder and autorenewal system, and to warn of any logistical or technological problems which may occur at larger-scale implementation.

### Methods for additional analyses (e.g. subgroup analyses) {20b}

The timing of the reminders relative to expiration date creates a staggered increase of treatment intensity at the beginning of the trial, and a decreasing intensity towards the end of the trial. In order to account for varying treatment intensity, we control for whether the full or a partial treatment was received by the participant. For this, we conduct sensitivity analyses which include binary and categorical variables in the logistic regression which denote the treatment intensity or variation received by a participant. This allows us to estimate an adjusted treatment effect, and to assess the marginal impact of an additional reminder message. In addition to the full and partial treatment effects, we will also estimate treatment effects for different sociodemographic subgroups using multiple logistic regressions controlling for gender, age group, and payment category.

### Methods in analysis to handle protocol non-adherence and any statistical methods to handle missing data {20c}

Non-adherence may result from individuals opting out of receiving messages and no longer receiving treatment. To assess the magnitude of non-adherence, we will compare opt-out rates between treatment arms. If opt-outs occur at high or at unequal rates between different treatment arms, an instrumental variable approach which instrumentalizes allocation for realized treatment status will allow for the estimation of an accurate local average treatment effect (LATE) despite non-adherence or technical failures [36]. Another potential source of non-adherence may stem from an autorenewal user from treatment arm A adding an individual from arm R or C to their renewal plan, resulting in a different treatment being administered. Therefore, autorenewal registrations across the different treatment arms will also be monitored. If a non-negligible number of participants is re-assigned thusly, the instrumental variable approach may also be used to account for this type of non-adherence.

Missing data is unlikely to occur as the routine data capture system usually ensures that all relevant routine data is collected automatically. If observations for time-consistent variables are missing from the data, they may be imputed from previous transaction records of an individual. Other missing data will not be imputed.

### Plans to give access to the full protocol, participant level-data and statistical code {31c}

The full protocol may be requested from the corresponding author anytime. Statistical code and survey data may be shared in approved cases. Routine data cannot be made available due to confidentiality restrictions.

### Oversight and monitoring

#### Composition of the coordinating centre and trial steering committee {5d}

The trial is coordinated by the Kwame Nkrumah University of Science and Technology (KNUST) School of Public Health (SPH) in collaboration with the Technische Universität Berlin (TUB) and the Ghana National Health Insurance Authority (NHIA). mTOMADY gGmbH is providing support for the development of the intervention, and formative research for intervention development. The two coordinating principal investigators (DO and WQ), together with the lead statistician (MS) have set up a Trial Management Committee, including representatives of the KNUST, TUB, and mTOMADY, which has regular monthly meetings. Trial data management is under the responsibility of KNUST. NHIA provides data to KNUST, which enables data access also to researchers from TUB. Local trial supervision is assured by senior KNUST staff and NHIA officials.

#### Composition of the data monitoring committee, its role and reporting structure {21a}

All routine data is monitored internally by the NHIA based on existing internal data and safety monitoring structures. Survey data is monitored at the KNUST by university data administrators independent of the trial research team. All data is curated by KNUST and it is only passed onto researchers of the consortium after all identifying information likely to compromise anonymity, privacy and confidentiality has been removed. The sponsor is independent and has no influence on the trial and its analysis.

#### Adverse event reporting and harms {22}

There will be no formal assessment or reporting of adverse events. However, opt-out responses will be monitored across all treatment arms. In case of high opt-out rates, interventions may be adjusted if deemed necessary by the management committee. The project team holds monthly meetings throughout the intervention period, where any unforeseen issues will be discussed and a response or possible adjustment to the protocol will be decided on by the management committee.

#### Frequency and plans for auditing trial conduct {23}

Researchers are monitoring intervention data at regular intervals during the intervention period. Emerging issues and potential problems will be discussed at the weekly meetings of the intervention team and during the monthly management committee meetings, where decisions might be taken on possible adjustments. The management committee is independent from the sponsor. Reporting requirements of the sponsor demand annual progress reports providing an update on project developments.

#### Plans for communicating important protocol amendments to relevant parties (e.g. trial participants, ethical committees) {25}

If necessary, required changes to the protocol will be communicated to the sponsor in the yearly progress report, to the ethical/review committees at both the NHIA and KNUST through amendment requests. Amendments will be transparently discussed in subsequent versions of the protocol and any resulting publications. Trial participants would be informed if protocol amendments would have relevant implications for their insurance renewal.

#### Dissemination plans {31a}

Trial results will be made publicly available and submitted to a relevant and peer-reviewed journal. Dissemination plans include several publications on the different aspects of the trial (effectiveness, usability, determinants of use, scale-up potential etc.). Sponsor regulations require open access publications. Results will also be presented at international and domestic conferences. In addition, results will be disseminated to NHIA officials, and together organize workshop presentations with other relevant stakeholders in Ghana including the Ministry of Health (MoH) and participating communities also with opinion leaders such as chiefs and community leaders. Also, evidence briefs will be produced on main findings and disseminated to relevant stakeholders in Ghana and other African countries, e.g. during the Conference on Public Health in Africa.

## Discussion

### Explanation of design choice

The phone numbers of members registered in the NHIS member database are often outdated. As a result, a renewal reminder sent to a number in the data base may not reach the intended recipient. Furthermore, an SMS message may reach an unintended recipient, because members sometimes register with a facilitators phone number (such as a local kiosk owner, NHIS staff, or medical staff), because phone numbers are passed on to friends or relatives, or because phone numbers are re-assigned by the telecommunications company after a period of inactivity. For a subset of 65,000 Kumasi residents, a phone number verification exercise was conducted to assess the magnitude of this problem. For all surveyed members, an up-to-date primary phone number was recorded for each individual. This exercise showed that only 48% of numbers in the data base matched the primary phone number stated by the member. Using this sample of recently verified phone numbers allows us to recruit a group of trial participants with a high likelihood of an SMS message reaching the intended recipient instead of another person (who may be in a different arm of the trial). This will allow for an evaluation with a high internal validity and result in an accurate treatment effect.

However, this type of phone number verification exercise is very resource-intensive and may not be scalable to the entire population. Furthermore, the verified number sample was not representative of the member population in Kumasi, as older and female members were more likely to be found in the survey locations during the verification exercise. Therefore, we also draw a random, representative sample from the NHIS data base to test the same intervention at a larger scale in a more realistic setting (excluding all those with phone numbers which were recorded in verified group). This will allow for an evaluation with a high external validity and an intent-to-treat effect. It will also allow us to assess the value of additional information and the necessity of such verification exercises. By combining the two approaches, we are able to evaluate both the effectiveness of the intervention and the potential for scale-up, without having to sacrifice internal or external validity. In the verified number sample, the differently sized treatment arms result from the fact that insurance status and expiration dates are not distributed equally throughout the sample. As a result, Group 1 is the largest, followed by Group 2, while Group 3 is very small. For the unverified number sample, we are able to randomly select individuals from a much larger pool of members in the NHIS database. Here, we chose to randomly draw equally sized Groups 1-3, which will result in equally sized treatment arms.

Due to the long run-time of the full treatment protocol (90 days for autorenewal, 120 days for reminders before and after expiration date), not all individuals will receive the full treatment. Individuals with expiration dates less than 30 (7, 1) days before trial start will not receive the t-30 (t_-7_, t_-1_) day reminder. Individuals whose expiration date is less than 60 (30) days away from the end of the intervention will not receive the t_+60_ (t_+30_) reminders. Furthermore, as individuals only receive the autorenewal savings reminders by signing up for the autorenewal system, those with expiration dates less than 90 or 60 days after sign-up will not receive the t_-90_ or t_-60_ savings reminder. We therefore expect that the aggregate treatment effect for the full trial sample will underestimate the actual treatment effect, as some individuals in the trial will not have received all messages due to these timing constraints. This will be addressed through subgroup and sensitivity analyses, which compares the outcomes for those who received the full treatment or a variation of a shortened treatment.

## Abbreviations

BMBF: Bundesministerium für Bildung und Forschung (German Ministry for Education and Research)

CIOMS: Council for International Organizations of Medical Sciences DHTF - Digital technologies for health financing

HHS: US Department of Health and Human Services

KNUST: Kwame Nkrumah University of Science and Technology

LATE: Local average treatment effect

MOH: Ministry of Health

NHIA: National Health Insurance Authority, Ghana

NHIS: National Health Insurance Scheme, Ghana

SMS: Short message service

TUB: Technische Universität Berlin

USSD: Unstructured Supplementary Service Data (“quick codes”)

WHO: World Health Organization

## Declarations

N/A

## Data Availability

NHIA routine data are confidential. Requests can be sent to NHIA, but decisions about sharing the data are solely at their discretion. Anonymized survey data can be made available upon request.

## Acknowledgements

We would like to thank our collaborators for their support: The NHIA of Ghana (especially Former CEO Dr. Lydia Dsane-Selby, Former Director Administration Dr. Francis-Xavier Andoh-Adjei, Former Director Research Raphael Segkpeb and Prof. Christian Agyare), the Ghana Ministry of Health (especially Head M&E Dr. Eric Nsiah-Boateng and Head Policy Mr. Benjamin Nyakutsey), Emergent Africa Payments, and MTN Ghana (especially Anita Obeng-Nkansah and Emmanuel Kojo Lartey).

## Authors’ contributions {31b}

All authors contributed to the conceptualization of the project. LN, MK, KM, DO, EO, CP, VS, WQ, and MS developed the methodology. EA, JA, OA, VA, DB, JC, JE, SK, LN, BO, DO designed and developed the intervention software. LN, EA, FA, JA, OA, VA, DB, FI, SK, BO, DO, EO, and MS acquired, validated, and maintained data. LN, EA, FA, JA, OA, VA, DB, FI, MK, KM, AN, BO, DO, EO, CP, MS, VS and RW conducted formal analyses and investigations for the study. EA, FA, JA, OA, VA, DB, SK, KM, BO, DO, EO and MS provided resources. EA, FA, JA, OA, VA, JB, DB, JE, KM, BO, DO and WQ, administrated the project. VA, JE, SK, KM, LN, BO, DO, EO, WQ and MS acquired funding. KM, DO, EO, WQ, MS, VS supervised research. LN wrote the original manuscript, WQ contributed. All authors critically reviewed and approved the final manuscript.

## Ethics approval and consent to participate {24}

This study was approved by the Kwame Nkrumah University of Science and Technology (KNUST) Committee on Human Research, Publication and Ethics. Written, informed consent will only be obtained from survey participants. No informed consent will be obtained from other participants.

## Consent for publication {32}

N/A

## Authors’ information

LN is first author, MS, WQ and DO are last co-authors. Correspondence to LN or to the principal investigator, WQ.

